# The correlation between small papillary thyroid cancers and gamma radionuclides Cs-137, Th-232, U-238 and K-40 using spatially-explicit, register-based methods

**DOI:** 10.1101/2023.01.11.23284363

**Authors:** Haytham Bayadsi, Paul Van Den Brink, Mårten Erlandsson, Soffia Gudbjornsdottir, Samy Sebraoui, Sofi Koorem, Pär Nordin, Joakim Hennings, Oskar Englund

## Abstract

A steep increase of small papillary thyroid cancers (sPTCs) has been observed globally. A major risk factor for developing PTC is ionizing radiation. The aim of this study is to investigate whether geological differences in the prevalence of sPTCs in Sweden are correlated to the deposit of Caecium-137, Thorium-232 (Th-232), Uranium-238 (U-238) or Potassium-40 (K-40) using different Geographical Information System (GIS) methods. Datasets of 812 sPTC patients were combined with the datasets of the total population in Sweden and were layered with the gamma radionuclide deposits. The prevalence of metastatic sPTC was associated with significantly higher levels of Gamma radiation from Th-232, U-238 and K-40. The observed results clearly indicate that sPTC has causative factors that are neither evenly distributed among the population, nor geographically, calling for further studies with bigger cohorts where environmental factors are believed to play a major role in the pathogenesis of the disease.

## 1. Introduction

Thyroid cancer (TC) is the most common form of cancer in the endocrine system, with papillary thyroid cancer (PTC) being its most common subtype (1). The incidence of PTC has been increasing steeply over recent decades, especially that of small papillary thyroid cancers (sPTCs) (≤ 20mm in size) and papillary thyroid microcarcinomas (PTCMs) (≤ 10mm in size). These are responsible for the greatest increase in incidence rate (2, 3). This increase can be only partially explained by diagnostic advances, such as the increased use of thyroid gland ultrasound (4). One etiologically established culprit is internal and external exposure to ionizing radiation, which is considered a major risk factor for TC in general and PTC specifically (5, 6).

Various studies have shown an increment in PTC cases in the areas exposed to nuclear reactor or weapons explosions, or populations exposed to external beam radiation to the head and neck areas, especially at a young age (7-11). Increased PTC risk has also been observed in survivors of the atomic bombs in Nagasaki and Hiroshima (12, 13). A study based on autopsy reports by Hayashi et al. (14) reported an increased prevalence of PTMC in the population of atomic bomb survivors in Japan in 1945, relating PTMC to radiation exposure. One of the most renowned examples is the Chernobyl nuclear reactor accident in 1986, resulting in thousands of new PTC cases in the population of children and adolescents at that time who were exposed to substantial doses of radiation (15). However, much larger populations in other countries were exposed to the nuclear fallout following the accident, Sweden being one of those (16). In Sweden, the largest fallout of radioactive substances occurred in parts of the eastern coast: one area measured ten times the normal level directly following the incident (17). The highest levels of Caesium 137 (Cs-137) activity exceeded 85 kBq/m^2^ and in some spots up to 200 kBq/m^2^ (18). Even though most areas in Sweden received very low radiation doses as a result of the fallout, an Uppsala University study showed a 5-fold higher overall cancer risk development for all cancer forms including PTC in the population with the highest exposure to the radioactive Cs-137 in Sweden (19). Cs-137 is a radioactive isotope of Caesium, produced by nuclear fission for use in medical devices and gauges. It is a biproduct of the nuclear fission process in nuclear reactors and weapons (20). The radiation that exists in Sweden today after the Chernobyl accident is mainly due to the 30-year long half-life of Cs-137 (17).

Aside from these specific exposure scenarios, the general population is also exposed to background radiation from Thorium-232 (Th-232), Uranium-238 (U-238) and Potassium-40 (K-40). Th-232 is a naturally occurring radioactive metal found in bedrock, soil, and water (21). It can be used to make ceramics, gas lantern mantles, welding rods and fuel for generating nuclear power as an alternative to uranium. Th-232 has been linked to an increased risk of developing liver, gallbladder and blood cancer and is classified by the International Agency for Research on Cancer (IARC) as carcinogenic to humans (22). To the best of our knowledge, there are no previous studies investigating the association between Th-232 levels and PTC incidence (21, 22).

U-238 is a naturally occurring radioactive element with a wide distribution in soil and higher concentrations in certain rock formations and water (9). Uranium has 17 known isotopes but only U-234, U-235, and U-238 are found in the environment due to their long half-lives (250 thousand, 700 million and 4.5 billion years respectively) (23). U-238 is released into the environment through wind and water erosion, as well as mining, milling or other forms of uranium processing (9). U-238 can be absorbed into the body through ingestion of contaminated food or drink, or inhalation of uranium-containing dust particles or aerosols (23). All uranium isotopes are radioactive, emitting alpha particles causing radiotoxicity (23). Therefore, chemical toxicity is the limiting factor determining the current US Environmental Protection Agency (EPA) maximum allowable contaminant level of 30 μg/L in drinking water (9). Increased thyroid cancer incidence has been documented in volcanic regions where increased urinary uranium concentrations have been found, although the underlying mechanism is still not clearly understood (9).

K-40 is an unstable and radioactive naturally occurring isotope of potassium. It has a very long half-life of 1.25 billion years (24) and makes up about 0.012% of the total amount of potassium found in nature. The radioactive decay used to calculate the potassium content is the gamma radiation from the decay of K-40 into argon-40. This decay branch represents 10.7% of the decay (25). After Th-232 and U-238, K-40 ranks 3^rd^ as a source of natural radioactivity contributing to earth heat (radiogenic heat) (24, 26). K-40 is the largest source of natural radioactivity in animals, including humans, because its traces are found in all potassium (24). A 70 kg human body contains 0.0164 grams of K-40, equalling an annual effective dose of 0.165 mSv, as compared to the estimated total effective dose from natural radionuclides in a human body of 0.3 mSv (27).

Geographic Information Systems (GIS) is a collective term for software that helps us capture, analyse, and visualize geographical information (28, 29). Maps allow us to show, understand, interpret, and visualize data in ways that can reveal relationships, patterns, trends, and geographical deviations. GIS gives us the opportunity to explore environmental risk factors (30) and to handle data covering big populations. This enables large scale geospatial assessments, which can be used to compute geostatistical correlations between disease prevalence and many potential environmental risk factors. GIS is primarily used in urban planning and social science and has not been yet used to any great extent in medical epidemiological research (31). This method is innovative and could draw attention to as yet unknown factors influencing thyroid cancer, and may potentially prove useful within other fields of medical research.

Unexplained regional differences in the distribution and increase of sPTC have been listed in Sweden (32). Early detection of more aggressive forms of thyroid cancer is essential for proper surgical and oncological treatment of these patients. The main aim of this study is to investigate whether geological differences in sPTC prevalence in Sweden are correlated to geographical factors in the surrounding environment such as the deposit of Cs-137 fallout following the Chernobyl nuclear powerplant accident, or the levels of the naturally occurring radioactive materials Th-232, U-238 or K-40.

## 2. Materials and methods

### 2.1 Patient selection

The study population originates from a nationwide cohort of patients with sPTC, all of whom were registered in the validated and prospectively maintained Scandinavian Quality Register for Thyroid, Parathyroid and Adrenal Surgery (SQRTPA) between 2011 and 2015. The SQRTPA national register was established in 2004 and is the world’s first quality register for endocrine surgery. It covers almost 100% of thyroid surgeries in Sweden. It is validated against the National Patient Register and is one of the few registers to have an internal quality audit that randomly checks the operating centres every year (33).

The inclusion criteria comprised a primary diagnosis of sPTC, defined as T1 (tumour ≤ 20mm). These T1 cases were then divided into subgroups depending on the lymph node status. The Nx group included patients where it was not possible to assess six or more lymph nodes. The N0 group included patients with no lymph node metastases, and the N1 group included patients with regional lymph node metastases indicating a more advanced disease. The inclusion was based on the Tumour–Node–Metastasis classification (TNM) 8th edition (34). Nx-classified tumours are clinically and prognostically treated as N0 groups and therefore were included in the N0 group. The Chi-squared test was used to compare the difference between genders. The Mann-Whitney U test was used to compare differences in age and tumour size. The statistical analyses were performed using SPSS statistics version 28 (IBM Corporate, Armonk, NY, USA).

Of the 14,827 thyroid surgeries that were performed during the study period, 933 patients were identified according to the inclusion criteria. Twenty-seven were excluded because they had tumours larger than 20mm (T-stadium > 1 as defined in the inclusion criteria) at the first surgery or at a complementary surgery. Eighty patients were excluded since we were unable to find their residential addresses and an additional 13 patients were excluded due to double registration. One patient was lost during data collection, leaving 812 patients to be included in the study (Table 1).

**Table 1.** Patient characteristics

|  | N1 | N0<br>(including Nx) | Total | P-value |
| --- | --- | --- | --- | --- |
| **Total number of patients** | 186 (23%) | 626 (77%) | 812 (100%) |  |
| **Sex** |  |  |  |  |
| Male | 43 (34%) | 106 (17%) | 149 (18%) |  |
| Female | 143 (66%) | 520 (83%) | 663 (82%) |  |
| Male/female ratio | 1:3.3 | 1:4.9 | 1:4.5 | 0.066 |
| **Age (years)** |  |  |  |  |
| Mean | 43.3 | 48.4 | 47.3 | <0.001 |
| Median | 42 | 49 | 47 |  |
| Range | 17-68 | 16-81 | 12-84 |  |
| **Tumour size (mm)** |  |  |  |  |
| Mean | 10.7 | 8.2 | 8.9 | <0.001 |
| Median | 11 | 7 | 8 |  |
| Range | 1-20 | 1-20 | 1-20 |  |

### 2.2 Spatial data

Three sources of natural gamma radiation were studied: Uranium-238 (U-238), Thorium-232 (Th-232) and Potassium-40 (K-40). To enable spatially explicit quantifications, we used national datasets of concentrations of U-238 (ppm eU), Th-232 (ppm eTh), and K-40 (%), respectively, with 200m resolution (Fig. 1), based on gamma spectrometric measurements from airplanes at low altitude (30-60m) (35).

**Figure 1.**
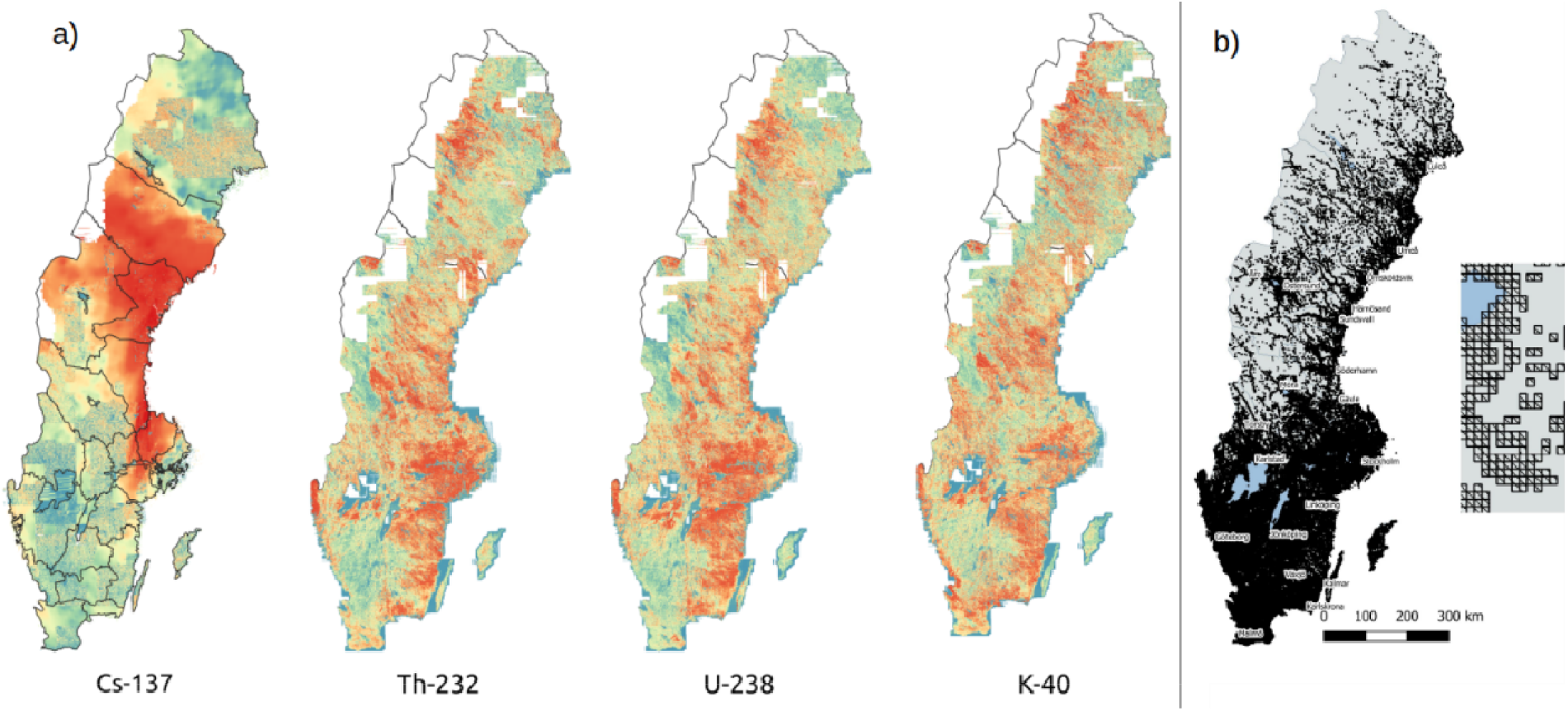
(a) Radiation levels displayed as percentiles for the respective datasets. Blue indicates relatively low levels and red indicates relatively high levels. Note that the maps are not comparable in terms of absolute radiation levels. (b) Population dataset used in the study. Black cells indicate inhabited areas.

Additionally, we studied the deposit of Caesium-137 (Ca-137) fallout after the Chernobyl 1986 incident. For this purpose, we used a national 200m dataset of estimated Cs-137 deposits on the ground in May 1986 (kBq/m2), based on recurring radiation measurements by airplanes done by the Swedish Radiation Safety Agency (SSM).

Population data from December 31^st^, 2019, were retrieved from the Swedish Tax Agency (Skatteverket) and Statistics Sweden (SCB), with two different resolutions, 1000m (Fig. 1) and 100m (36).

### 2.3 Spatial analysis

The analysis was divided into two parts. First, we conducted a geospatial analysis of the distribution of cases across Sweden, to assess the extent to which prevalence can be predicted by population and to identify potential areas where sPTC is over- or under-represented. Second, we conducted a statistical analysis, where we assessed the degree to which the patients may have been subject to different levels of radiation compared with the total population.

#### 2.3.1 Spatial distribution of cases

Every person who lives in Sweden receives a ten-digit Swedish personal identity number (PIN). The Swedish PIN covers almost 100% of the Swedish health care system and can be a useful link between medical registers (37). Residential addresses were retrieved through the Swedish Tax Agency (Skatteverket) and Statistics Sweden (SCB) crosslinking pseudonymised data from the SQRTPA. Google’s Application Program Interface (API) was then used to convert the addresses of the 812 patients to geographical coordinates.

Several methods were then tested to assess the distribution of cases, as follows:

##### 2.3.1.1 Aggregated prevalence

The number of cases per 10,000 inhabitants was calculated for multiple geographical areas, of which two are presented here. First, the GIS software QGIS (38) was used to count the number of cases in each county and the total population, using an official county polygon dataset. The field calculator was then used to calculate the corresponding prevalence. The same was calculated for each cell in the 1000m population dataset using the GIS software GRASS GIS (tool: v.vect.stats). As a comparison, the expected number of cases in each cell was calculated based on the population in each cell and the national prevalence. The latter was calculated based on the total population as defined by the same population dataset.

##### 2.3.1.2 Heatmaps and spatial distribution compared to population

Heatmaps can be used to identify “hotspots” within a geographical area, and thus can be potentially useful for indicating areas where sPTC is over- or under-represented. For this purpose, we used the Kernel Density Estimation tool in QGIS with standard quartic kernel shape, a pixel size of 100m, and a radius of 20,000m. First, two approaches were tested to indicate hotspots, or over-representation of sPTC, with the “weight from field” set to (a) prevalence within a 1000m buffer from each sPTC patient and (b) prevalence within a circle that extends to the nearest other sPTC patient. Then, an attempt was made to identify areas where sPTC is under-represented, with the “weight from field” set to the population within a circle that extends to the next nearest sPTC patient multiplied by radius of the circle.

To test the degree to which prevalence is connected to population density, we first divided all population cells into deciles (based on population values). We then calculated the number of patients residing in population cells belonging to the respective deciles. Finally, we conducted 25 simulations, where 812 cases were randomly assigned to population cells based on the rule that the probability of a case is directly proportionate to the population density. That is, the probability of assigning a case to a cell with population value 1,000 is 100 times greater than the probability of assigning a case to a cell with population value 10. We then counted the number of simulated cases in population cells belonging to the previously identified deciles in each of the 25 simulations, and calculated the resulting median, 2^nd^ quartile, and 3^rd^ quartile number of simulated cases.

##### 2.3.1.3 Prevalence among neighbours to patients

In addition to heatmaps, spatial patterns in prevalence were assessed using a k-nearest-neighbour (KNN) approach. First, a national grid with a resolution of 100m was created and the population and number of patients were calculated for each cell. Second, a circle around each cell was expanded until it contained 10,000 and 50,000 people, respectively. Third, based on the number of cases in the circle, the prevalence of sPTC among the 10,000 and 50,000 nearest neighbours to each cell was calculated, respectively. The KNN analyses were performed using the software Equipop (39).

#### 2.3.2 Statistical analysis

##### 2.3.2.1 Comparison of sPTC patients and the entire Swedish population in terms of gamma radiation levels at the location of residence

To identify differences between the patients and the total population, in terms of gamma radiation levels at the location of residence, we first assigned radiation values for Cs-137, U-238, Th-232, and K-40, for each patient (using GRASS GIS: w.what.rast), using point data for patients and the different radiation raster datasets. The output was saved as a csv file, for further statistical analysis using a stand-alone Python script, as described below. The same procedure was then repeated using the 100m population vector dataset instead of patient point data.

The csv data for the cases consisted of a table with one row per patient and multiple columns for patient information, including whether sPTC was metastatic or not, and radiation levels for the assessed radiation sources. Population data, however, consisted of a frequency table with one row for each 100m population square, and columns for total population, radiation levels, and the number of cases subdivided into metastatic and non-metastatic disease. To facilitate a statistical comparison between the sample and the population, the population table had to be converted to a raw dataset with one individual per line. This was done using Python.

To illustrate the distribution of radiation levels for the patients and the population, respectively, box plots were created using the Pandas boxplot function (whiskers showing the 10^th^ and 90^th^ percentiles). Two-sample t-tests were then conducted for the sample and population datasets to assess how their respective means differed. The ratio of variance (patients: population) was calculated and considered acceptable (0.59 for Cs-137 and near 1.0 for other radiation sources).

##### 2.3.2.2 Radiation patterns among nearest neighbours

For each 100m grid cell (see 2.3.1.3), the average radiation value was extracted from the four radiation maps (Fig. 1). We used statistical modelling to explore the patterns between the number of cases among the 10,000 and 50,000 nearest neighbours to each cell, respectively, and the four radiation variables. Because of non-linear patterns in the data, generalized additive models (GAM) were used via the R package mgcv (40).

## 3. Results

### 3.1 Spatial prevalence

The 812 cases of sPTC do not seem to be geographically distributed solely according to population density. While following the population density fairly well, they are over-represented in cells (1000m) that are densely populated and under-represented in sparsely populated cells (Fig. 2). The simulation, naturally, resulted in a distribution of cases very similar to the population density. This means that there should be explanatory variables other than population density in the direct vicinity.

**Figure 2.**
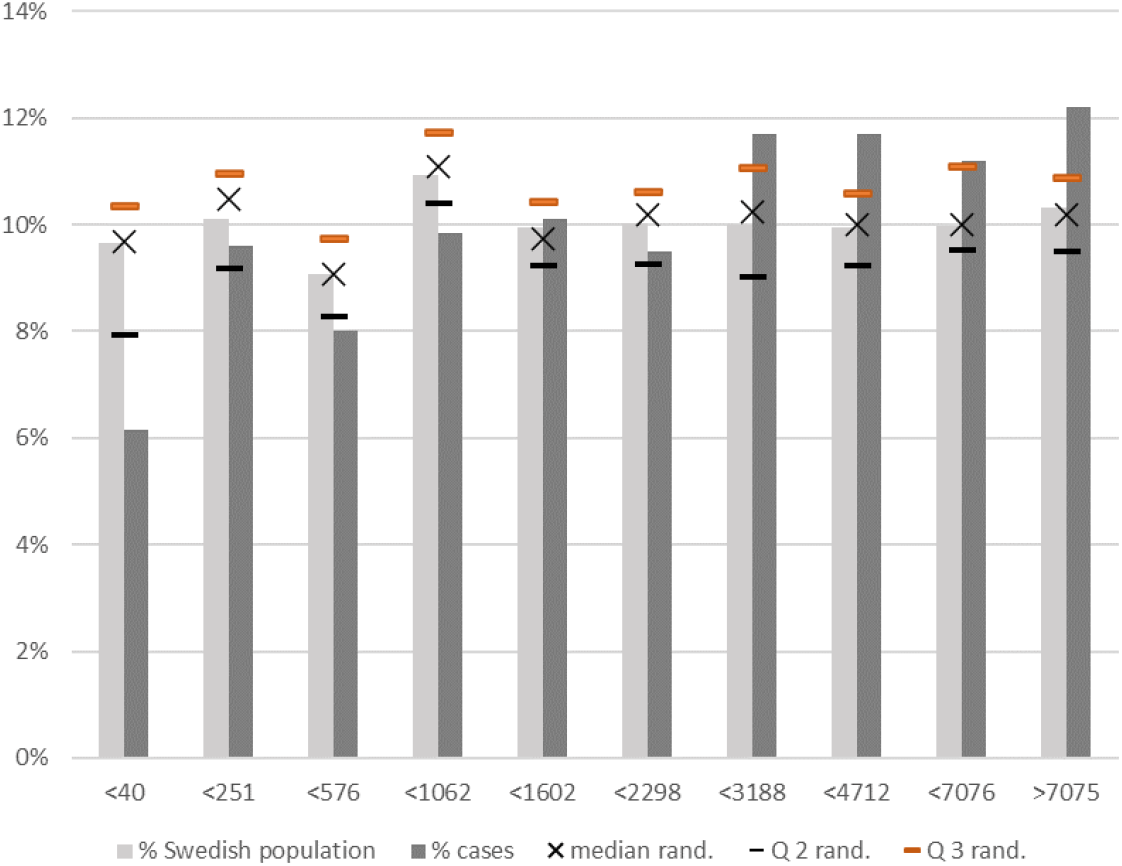
Share of (i) total population, (ii) sPTC cases, and (iii) simulated sPTC cases, where the probability of a case is directly proportionate to the population, divided into deciles of total population per cell (x-axis). “Median rand” refers to the median share of total simulated cases per cell in the respective population density intervals, and “Q2 rand” and “Q3 rand” refer to the corresponding second and third quartile, respectively.

At the county level, there are notable differences in prevalence: from 0.277 (Dalarna) to 1.153 (Jämtland) cases per 10,000 inhabitants (Fig. 3A; cf the national prevalence of ca. 0.81). The two heatmaps aimed to identify areas with an over-representation of sPTC cases: Figure 3, B and C show notable discrepancies and thus fail to provide a conclusive picture. The heatmap aimed at identifying areas with an under-representation of sPTC cases, but shows some similarities to the county aggregates. The sPTC cases are thus neither evenly distributed by location or by population density.

**Figure 3.**
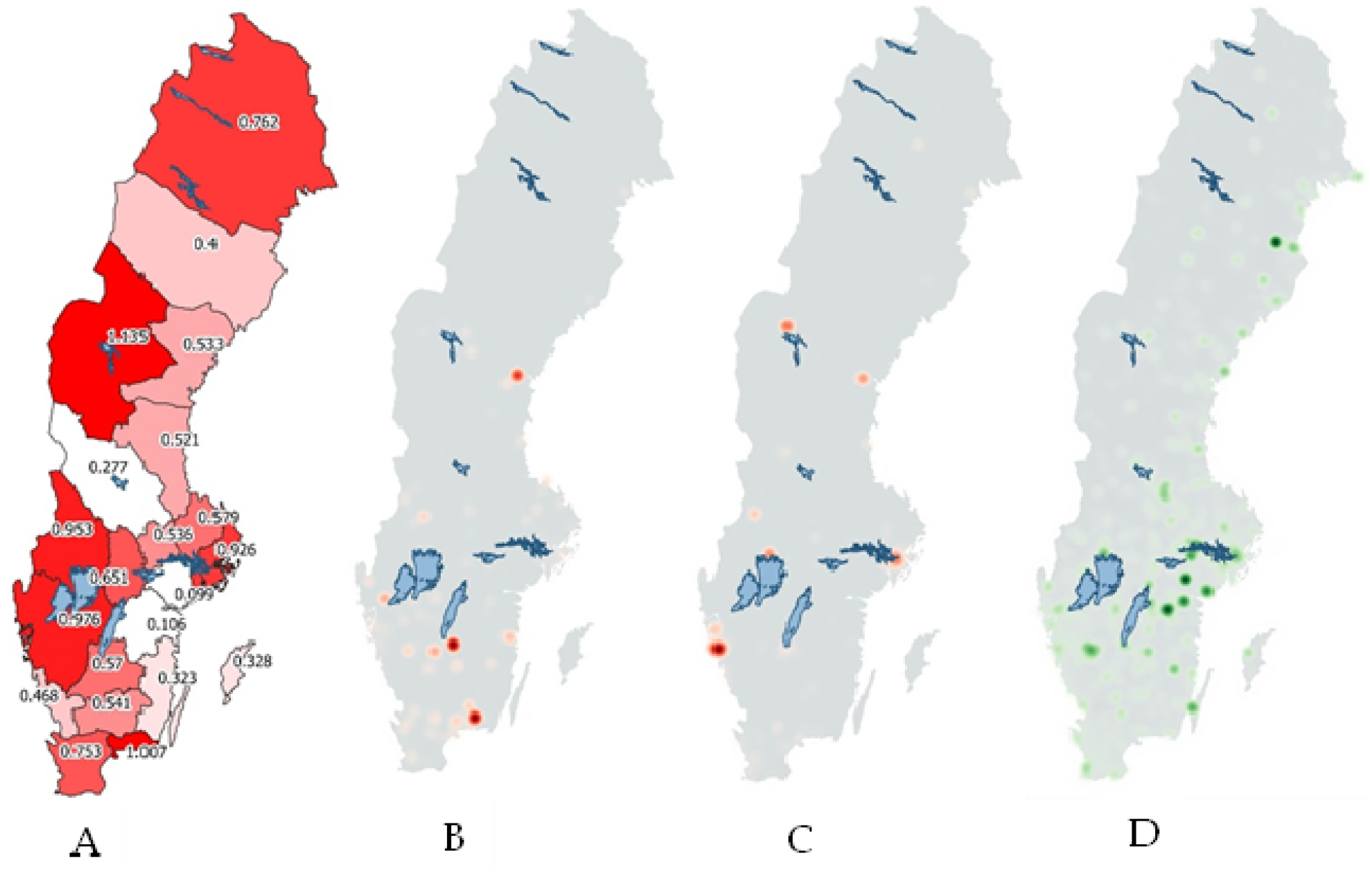
(A) The prevalence by county (number of cases per 10,000), (B) heatmap with a buffer of 1km around each case, (C) heatmap with a buffer to nearest neighbour case, showing where prevalence is higher compared to population within the buffer zones and (D) heatmap showing areas with fewer cases than expected based on distance to cases and population (green).

The analysis of prevalence among the nearest neighbours to each 100m cell reveals some additional detail, indicating that there is an over-representation of sPTC in densely populated areas near the major cities, but even in more rural areas, as shown with the red areas in Figure 4. Using the nearest 10,000 or 50,000 neighbours’ results in similar spatial patterns, the former results in a clearer delineation of hotspots than the latter.

**Figure 4.**
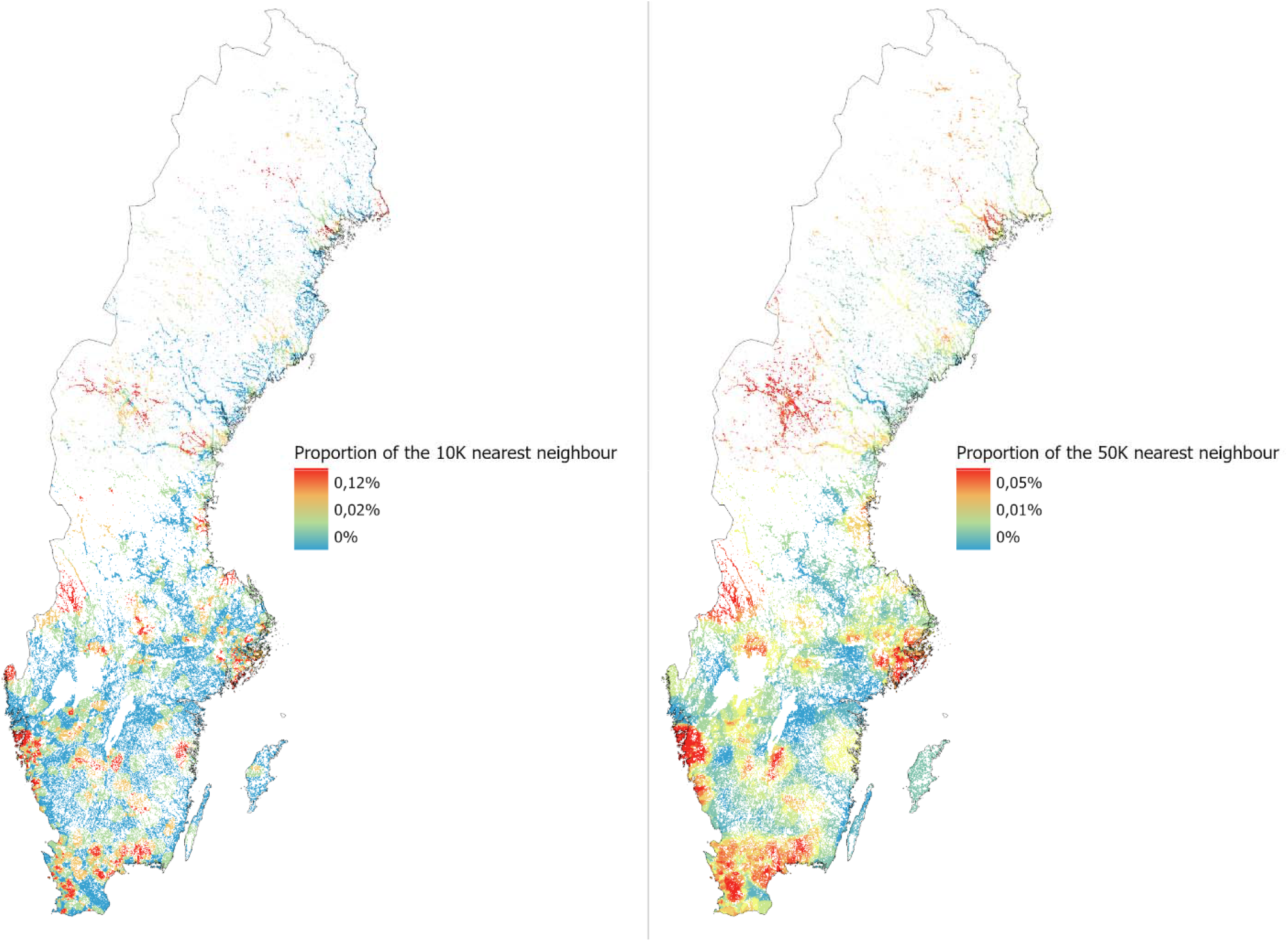
The prevalence among the 10,000 (left map) and 50,000 (right map) nearest neighbours to each 100m populated cell in Sweden.

### 3.2 Relationships between sPTC and gamma radiation

Differences between our study cohort and the entire population of Sweden in terms of radiation levels cannot be determined by visual assessment of boxplots (Fig. 5). There are, however, some notable differences between metastatic and non-metastatic patients in this respect, indicating that patients with metastatic sPTC are generally subject to higher levels of gamma radiation from U-238 and Th-232 and lower levels of radiation from Cs-137 (Fig. 5).

**Figure 5.**
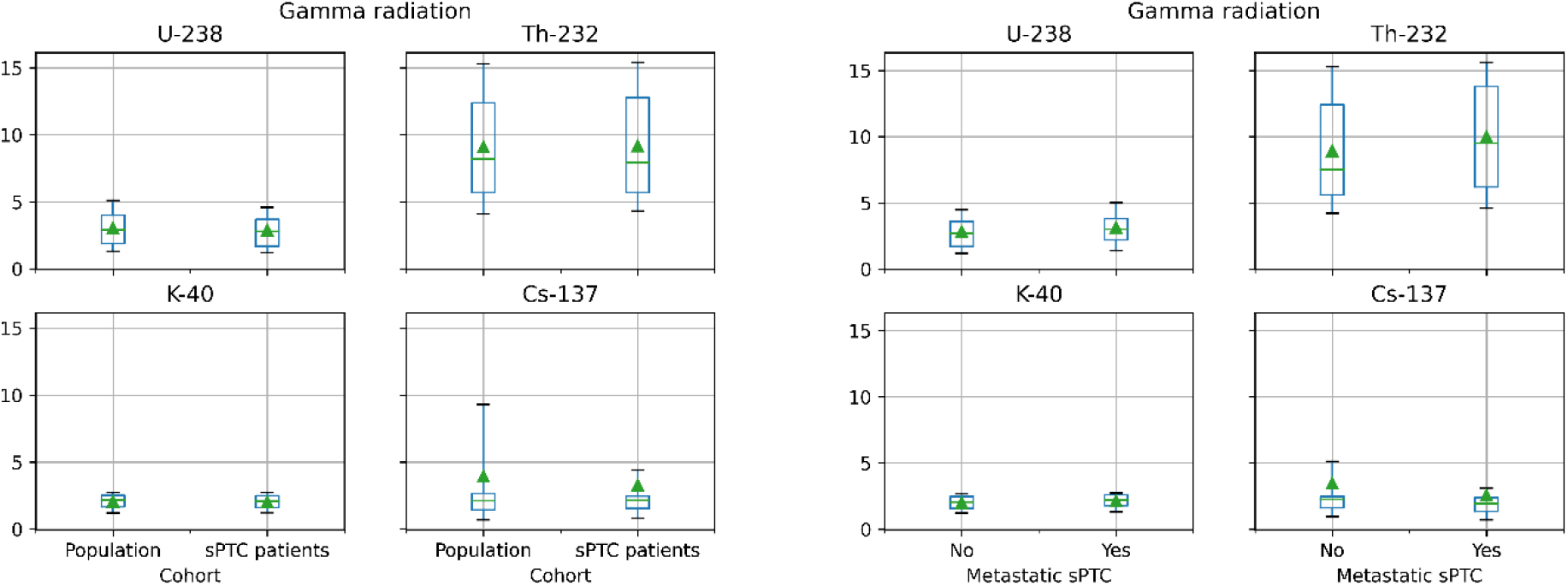
Whisker diagrams illustrating the statistical degree of gamma radiation from the assessed sources, for the patient sample and the entire Swedish population, respectively (left), and patients with metastatic and non-metastatic sPTC, respectively (right). Box: median, 1^st^ to 3^rd^ quartiles; Whiskers: 10^th^ and 90^th^ percentile; Triangle: mean; Units: U-238 (ppm eU), Th-232 (ppm eTh), K-40 (%), Cs-137 (kBq/m2). Metastatic sPTC refers to small thyroid papillary cancers with central and/or lateral lymph node metastasis, indicating a more advanced disease

The statistical analysis reveals some significant, and unexpected, differences between the groups (Table 2):

**Table 2.** Key statistical properties for gamma radiation levels for the assessed sources, for all cases, metastatic cases (N1), non-metastatic cases (N0), and the entire Swedish population.

|  |  | All cases | Metastatic cases (N1) | Non-metastatic cases (N0) | Entire population |
| --- | --- | --- | --- | --- | --- |
| **Caesium-137** | Mean (kBq/m2) | 3.28 | 2.57 | 3.49 | 3.95 |
|  | Std | 5.23 | 3.46 | 5.63 | 6.80 |
|  | Difference between patients and population | **Significantly lower (p < 0.01)** | **Significantly lower (p < 0.01)** | Insignificantly lower (p = 0.09) |  |
|  | Difference between metastatic and non-metastatic cases |  | **Significantly lower (p < 0.05)** |  |  |
| **Thorium-232** | Mean (ppm eTh) | 9.16 | 9.99 | 8.92 | 9.09 |
|  | Std | 4.40 | 4.37 | 4.38 | 4.35 |
|  | Difference between patients and population | Insignificantly higher (p = 0.64) | **Significantly higher (p < 0.01)** | Insignificantly lower (p = 0.31) |  |
|  | Difference between metastatic and non-metastatic cases |  | **Significantly higher (p < 0.01)** |  |  |
| **Uranium-238** | Mean (ppm eU) | 2.89 | 3.13 | 2.83 | 3.06 |
|  | Std | 1.54 | 1.37 | 1.59 | 1.61 |
|  | Difference between patients and population | **Significantly lower (p < 0.01)** | **Significantly higher (p < 0.001)** | Insignificantly lower (p = 0.57) |  |
|  | Difference between metastatic and non-metastatic cases |  | **Significantly higher (p < 0.05)** |  |  |
| **Potassium-40** | Mean (%) | 2.04 | 2.14 | 2.01 | 2.06 |
|  | Std | 0.60 | 0.56 | 0.60 | 0.61 |
|  | Difference between patients and population | Insignificantly lower (p = 0.35) | Insignificantly higher (p = 0.06) | **Significantly lower (p < 0.05)** |  |
|  | Difference between metastatic and non-metastatic cases |  | **Significantly higher (p < 0.01)** |  |  |

- sPTC patients, in general, are not subject to significantly higher levels of gamma radiation from any of the assessed sources, although they have significantly *lower* levels of gamma radiation from U-238 and Cs-137, compared to the Swedish population as a whole.
- Patients with non-metastatic sPTC are subject to significantly lower levels of gamma radiation from K-40, compared to the Swedish population as a whole.
- Patients with metastatic sPTC are subject to significantly higher levels of gamma radiation from Th-232 and U-238, but significantly lower levels of gamma radiation from Cs-137, compared to the Swedish population as a whole.
- Patients with metastatic sPTC are subject to significantly higher levels of gamma radiation from Th-232, U-238, and K-40, but significantly lower levels of gamma radiation from Cs-137, compared to patients with non-metastatic sPTC.

The statistical modelling of sPTC cases relative to different radiation levels among a set number of “nearest neighbours” to each sPTC patient is generally inconclusive (Fig. 6). There appears to be a greater risk of sPTC at certain levels of radiation from all the different sources, but there are no logical explanations for such “bumps”. The results also indicate that (Fig. 6a) very high concentrations of K-40 are associated with a greater risk of sPTC, (Fig. 6b) there is initially a linear increase in sPTC cases with increasing radiation levels from K-40 (which then levels out) and Th-232 (which is then reversed). These findings should, however, be interpreted with caution.

**Figure 6.**
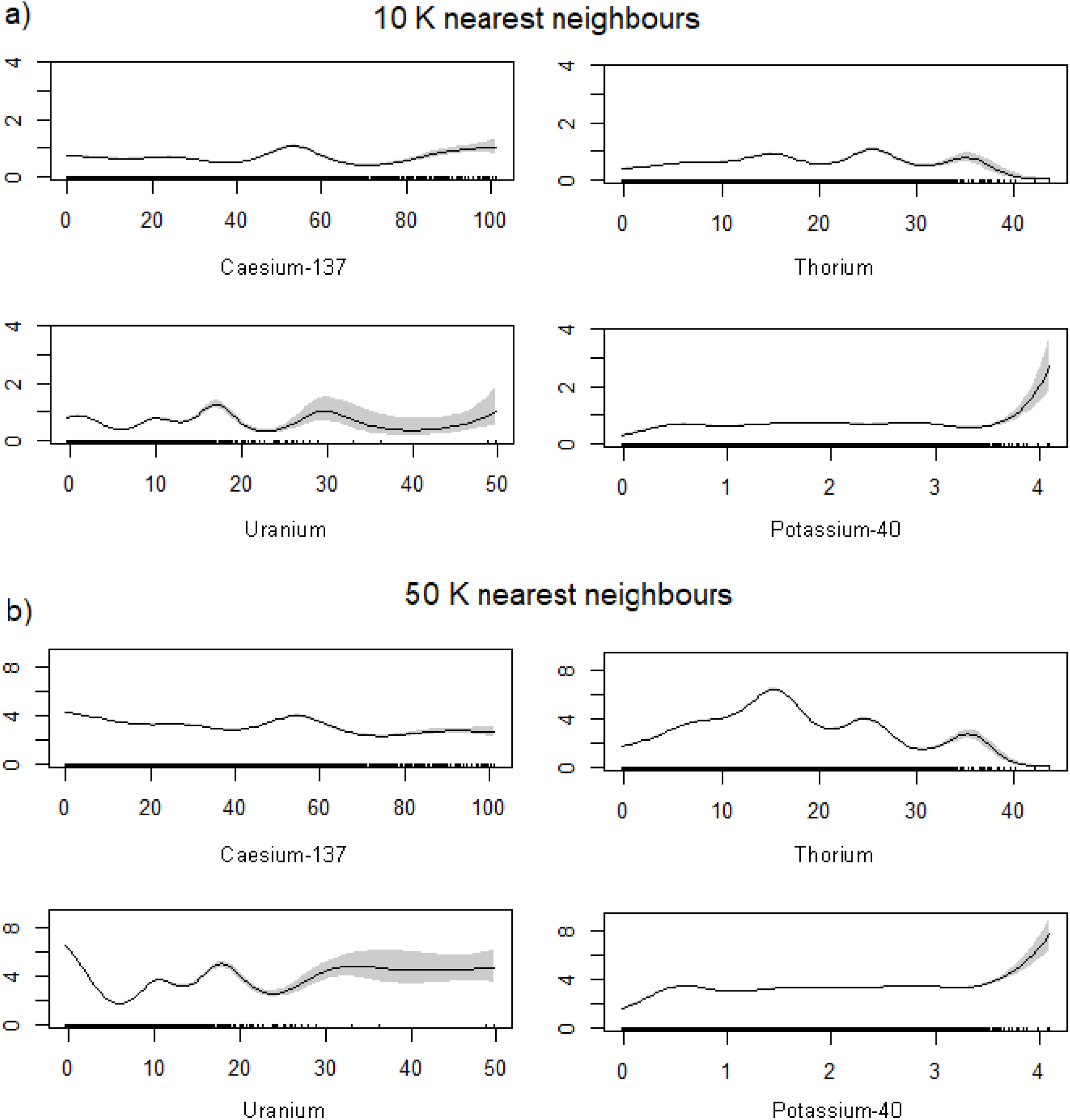
The partial response plots of the four radiation variables in the GAM models. a) shows the partial responses from the model using each cell’s 10,000 nearest neighbours and b) those with the 50,000 nearest neighbours. The x-axis shows the value of the radiation layer in each cell and the y-axis shows the modelled number of patients within the chosen number of nearest neighbours.

## 4. Discussion

The different approaches to identifying spatial distribution of sPTC prevalence show some common patterns, although additional studies with larger sample sizes are necessary to provide conclusive results. However, it is obvious that sPTC has causative factors that are neither evenly distributed among the population, nor geographically.

The results show that metastatic sPTCs are associated with significantly higher levels of gamma radiation from Th-232, U-238 and K-40, indicating that increased levels of gamma radiation from the environment could be a risk factor for developing metastatic sPTC. There is, however, no evidence to suggest that it could be a risk factor for developing non-metastatic sPTC. It was unexpected that sPTC patients, according to our analysis, are subject to lower levels of gamma radiation from Cs-137, but this may be explained by methodological limitations. While gamma radiation from U-238, Th-232, and K-40 can be considered relatively constant over time, the Cs-137 data we used in the study refer to levels at the time of disposal following the Chernobyl accident, which (given the relatively short half-life of Cs-137) may not be representative of current conditions. Future studies of connections between sPTC and Cs-137 should take such temporal aspects into consideration.

Spatially explicit register-based studies, such as shown here, are essential to studying any disease with unknown causative factors associated with the physical environment, or otherwise spatially dependent, and may thus have a notable potential for future research. One example where this kind of study could provide important scientific advances is Type 1 Diabetes Mellitus (DM), for which Sweden has the second highest incidence in the world. Since this cannot be explained by genetics, the possibility of environmental risk factors has been raised multiple times (41). Spatially explicit register-based studies can be used to eliminate or confirm suspected environmental risk factors but also, given the abundance of geographical data, can identify new causative factors associated with the physical environment, or new spatial patterns that can provide additional information for better understanding the cause of the disease. Another strength of this kind of study is that it allows us to use the entire population as a comparative cohort, without requiring any information about individuals.

Several additional methodological limitations may have affected the results. First, the analysis lacks information regarding where the patients were born and lived before the time of diagnosis. Some patients may have moved several times from birth to time of diagnosis, so the gamma radiation levels in the location where they lived at the time of diagnosis may not be representative of the gamma radiation they were subjected to during their childhood. Even though most of the population remains stable over time in a geographical region (42), there is a constant movement of people between regions. Furthermore, almost all people that remain in a region move within it at least once. Since there is a higher risk of developing PTC when exposed to ionizing radiation in childhood than in adulthood (7, 8), this is an important limitation of this study. Future studies should attempt to track the movement of the patients and aggregate radiation levels from different residence locations.

The limited sample size probably explains why the results are inconclusive and sometimes contradictory. Considering the geographical prevalence, this is particularly problematic in a country like Sweden, with large variations in population density across the country. One single case in a sparsely populated area could mean an over-representation compared to what can be expected based on the population in the area used for aggregation. It would therefore be misleading to report differences in prevalence for areas that are too small, in terms of population. A larger sample size would facilitate the use of smaller aggregation units but would also enable a better basis for geostatistical analysis. Considering radiation, or other potential risk factors that can be similarly assessed, a larger sample size would make the spatiotemporal concerns raised above less problematic. However, provided that the sample is truly random and that the spatiotemporal concerns can be properly addressed, the limited size of the sample should not be significant.

There are multiple challenges in determining suitable methods for assessing spatial differences in prevalence. Both heatmaps and maps based on prevalence among nearest neighbours are sensitive to sample size, and the location of very few cases in sparsely inhabited areas can cause large areas to be identified as “hot spots”. There are also other methodological concerns that should be noted. Heatmaps, for example, have been used in a few similar studies (43), but they are principally a kind of logical model (44) requiring careful attention to detail in design and parameterization, as well as empirical validation, to be useful. Both the former and the latter are difficult. However, if done appropriately, they are powerful in determining areas with an over- or under-representation, especially since they are not limited to a predefined aggregation unit, which may or may not be appropriate given underlying (unknown) causative factors. That is, models that are not dependent on predefined borders for aggregation may be more effective in identifying geographical patterns. Regardless of the method used to identify spatial differences in prevalence (for any disease), it is imperative to use appropriate methods and to validate (or at least evaluate) the results using empirical data. In many cases, it may be wise to use multiple methods that provide complementary information, such as demonstrated in our study.

Given the above discussion, further analysis of sPTC requires a larger sample size and more information about all addresses of all patients from their time of birth to their time of diagnosis.

## 5. Conclusions

Metastatic sPTCs are associated with significantly higher levels of gamma radiation from Th-232, U-238 and K-40, indicating that increased levels of gamma radiation from the environment could be a risk factor for developing metastatic sPTC. There is, however, no evidence for suggesting that it could be a risk factor for developing non-metastatic sPTC.

The results in this study clearly indicate that sPTC has causative factors that are neither evenly distributed among the population, nor geographically. Spatially dependent factors could be either demographic or environmental. Further analysis of sPTC requires a larger sample size and more information about all addresses of all patients from their time of birth to their time of diagnosis.

Spatially explicit register-based studies, such as shown here, are essential to studying any disease with unknown causative factors associated with the physical environment, or otherwise dependent on location, and may thus have significant potential for future research.

## Data Availability

All data produced in the present study are available upon reasonable request to the authors

## List of abbreviations

TC: Thyroid cancer
PTC: Papillary thyroid cancer
sPTC: Small papillary thyroid cancer
PTCM: Papillary thyroid microcarcinoma
GIS: Geographic Information System
QGIS: Quantum Geographic Information System
GRASS: Geographic Resources Analysis Support System
Cs-137: Caesium-137
Th-232: Thorium-232
U-238: Uranium-238
K-40: Potassium-40
SQRTPA: Scandinavian Quality Register for Thyroid, Parathyroid and Adrenal Surgery
API: Application Program Interface

## 6. Additional information

### Funding

This study was funded by the Unit of Research Education and Development, Jämtland-Härjedalen County, Sweden (Grant number: JLL-940567) and Jämtland County Cancer and Care Fund, Sweden (Grant number: 748). Open access funding provided by Umeå University, Sweden.

### Author contributions

H. Bayadsi, P. Van Den Brink, M. Erlandsson, P. Nordin, J. Hennings and O. Englund contributed to the study concept and design. Material preparation and data collection were performed by H. Bayadsi and S. Koorem. Data analysis was done by H. Bayadsi, P. Van Den Brink, M. Erlandsson and O. Englund. The first draft of the manuscript was written by H. Bayadsi, S. Gudbjornsson, S. Sebraoui and O. Englund. All authors commented on previous versions of the manuscript and read and approved the final manuscript.

### Competing interests

The authors declare that they have no conflicts of interest.

### Ethics

This study was approved by the Swedish Ethical Review Authority, permit number 2019-04983, and was carried out in accordance with the EU’s General Data Protection Regulation (GDPR) rules.

## Notes

### Competing Interest Statement

The authors have declared no competing interest.

### Funding Statement

This study was funded by the Unit of Research Education and Development, Jamtland-Harjedalen County, Sweden (Grant number: JLL-940567) and Jamtland County Cancer and Care Fund, Sweden (Grant number: 748). 
Open access funding provided by Umea University, Sweden.

### Author Declarations

This study was approved by the Swedish Ethical Review Authority, permit number 2019-04983, and was carried out in accordance with the EUs General Data Protection Regulation (GDPR) rules.

